# Machine Learning to Predict 10-year Cardiovascular Mortality from the Electrocardiogram: Analysis of the Third National Health and Nutrition Examination Survey (NHANES III)

**DOI:** 10.1101/2021.09.09.21263327

**Authors:** Chang H. Kim, Sadeer Al-Kindi, Yasir Tarabichi, Suril Gohel, Riddhi Vyas, Shankar Srinivasan

**Author notes:** **Corresponding Author:** Chang H. Kim, MD PhD, Division of Cardiovascular Medicine, Johns Hopkins Hospital, Address: 1800 Orleans St, Zayed 7122, Baltimore, MD 21287.

## Abstract

**Background:** The value of the electrocardiogram (ECG) for predicting long-term cardiovascular outcomes is not well defined. Machine learning methods are well suited for analysis of highly correlated data such as that from the ECG.

**Methods:** Using demographic, clinical, and 12-lead ECG data from the Third National Health and Nutrition Examination Survey (NHANES III), machine learning models were trained to predict 10-year cardiovascular mortality in ambulatory U.S. adults. Predictive performance of each model was assessed using area under receiver operating characteristic curve (AUROC), area under precision-recall curve (AUPRC), sensitivity, and specificity. These were compared to the 2013 American College of Cardiology/American Heart Association Pooled Cohort Equations (PCE).

**Results:** 7,067 study participants (mean age: 59.2 ± 13.4 years, female: 52.5%, white: 73.9%, black: 23.3%) were included. At 10 years of follow up, 338 (4.8%) had died from cardiac causes. Compared to the PCE (AUROC: 0.668, AUPRC: 0.125, sensitivity: 0.492, specificity: 0.859), machine learning models only required demographic and ECG data to achieve comparable performance: logistic regression (AUROC: 0.754, AUPRC: 0.141, sensitivity: 0.747, specificity: 0.759), neural network (AUROC: 0.764, AUPRC: 0.149, sensitivity: 0.722, specificity: 0.787), and ensemble model (AUROC: 0.695, AUPRC: 0.166, sensitivity: 0.468, specificity: 0.912). Additional clinical data did not improve the predictive performance of machine learning models. In variable importance analysis, important ECG features clustered in inferior and lateral leads.

**Conclusions:** Machine learning can be applied to demographic and ECG data to predict 10-year cardiovascular mortality in ambulatory adults, with potentially important implications for primary prevention.

## INTRODUCTION

Atherosclerotic cardiovascular disease (ASCVD) is a major cause of morbidity and mortality in the United States, with an incidence of 580,000 myocardial infarctions and 610,000 strokes occurring each year^1^. The current standard risk calculator in the U.S., the 2013 American College of Cardiology and American Heart Association Pooled Cohort Equations (PCE)^2^, utilizes demographic and clinical variables (age, sex, race (white or African American), total cholesterol, high-density lipoprotein cholesterol, systolic blood pressure, treatment for hypertension, diabetes mellitus, and smoking status) to estimate 10-year risk of incident ASCVD events, defined as fatal and nonfatal myocardial infarction or stroke. While the PCE is in routine clinical use in the U.S., it has been criticized for its suboptimal calibration and risk prediction in various patient populations^3^, and the requirement for blood draws represents a hindrance in its utility as a primary screening tool.

The electrocardiogram (ECG) is widely used in clinical practice to diagnose various cardiac conditions, such as myocardial infarctions, arrhythmia, and others. While prior studies have identified individual ECG components as predictors of adverse cardiovascular events^4-7^, the value of aggregate screening ECG data for prediction of long-term cardiovascular outcomes is less well understood. Given their highly correlated nature, ECG data are well suited for analysis by machine learning methods, and recent studies have found machine learning on ECG data to be useful in identifying specific cardiac disease states^8-12^. Considering such background, we sought to examine whether machine learning methods could predict 10-year cardiovascular mortality from aggregate ECG data in an ambulatory population.

## METHODS

### Data source

This study utilized the third iteration of the National Health and Nutrition Examination Survey (NHANES III)^13^, which consists of healthcare survey data compiled from a nationally representative sample of 39,695 persons from 1988 to 1994. In addition to demographic, historical, and physical items from the questionnaire and examination, biochemical laboratory studies and ECG data are available for a subset of the surveyed population. Mortality outcomes, including the cause of death, are available via linked National Death Index files^14^. Since NHANES III is a publicly available, de-identified data set, a separate Institutional Review Board review was not required for this study.

This study included NHANES III participants who were 18 and older whose demographic and mortality data were available, and who did not have pre-existing cardiovascular disease, defined as lack of self-reported history of myocardial infarction, congestive heart failure, or stroke. Among this group, 7,067 participants had additional data from a standard 12-lead ECG and cardiovascular risk factors required to compute the PCE, who formed the main study cohort for this analysis. The study flow diagram is shown in **Figure 1**.

**Figure 1.**
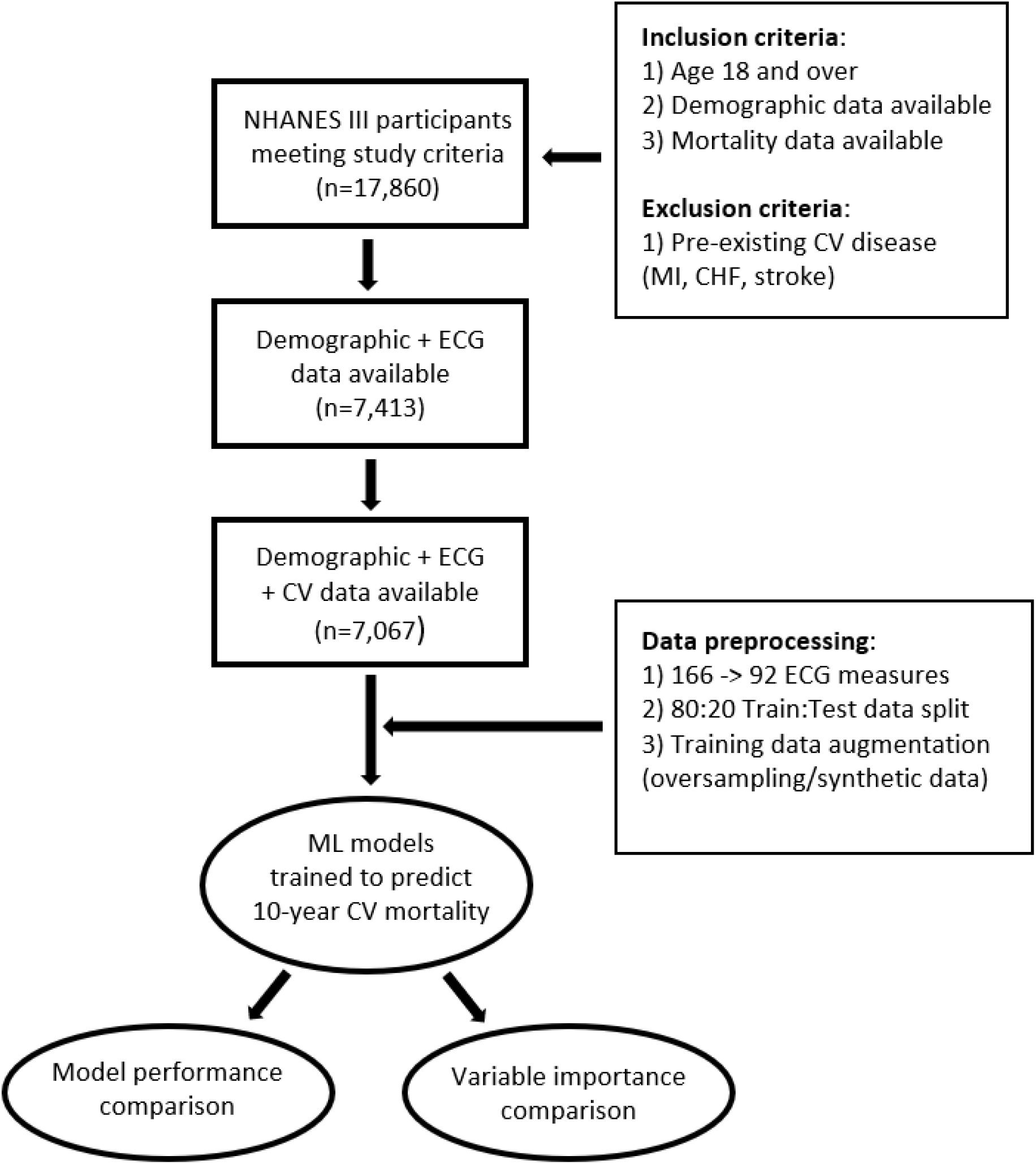
Study flow diagram CHF: congestive heart failure, CV: cardiovascular, ECG: electrocardiogram, MI: myocardial infarction, ML: machine learning, NHANES: National Health and Nutrition Examination Survey

### Data preparation

All data were imported and analyzed using R 3.5.1 statistical software^15^. Publicly available R packages were utilized for data preparation (*tidyverse*^16^, *mice*^17^, *ROSE*^18^), general machine learning (*caret*^19^), deep learning (*Keras*^20^), ensemble learning (*SuperLearner*^21^), classification performance evaluation (*precrec*^22^), and survival analysis and plotting (*survival*^23^, *survminer*^24^). Details of data preparation, model training, and performance evaluation are described below.

Demographic and clinical data were recorded from relevant sections of NHANES III. Baseline age and sex were recorded as reported at time of survey. Race categories were simplified to White, Black, or Other to avoid data sparsity. For vital signs, the median value of multiple measurements was recorded. For clinical data, an affirmative answer on the questionnaire or relevant medication and laboratory measurements were utilized (e.g. for history of diabetes, qualifying criteria included answering ‘yes’ to questionnaire or having laboratory values of any fasting glucose ≥126 mg/dL or hemoglobin A1c ≥ 6.5%). For participants taking medications for high cholesterol, laboratory values were adjusted to reflect average statin effect (total cholesterol: 21% reduction, high-density lipoprotein cholesterol: 3.5% increase)^25^.

For outcomes, death status and cause of death were determined using International Statistical Classification of Diseases and Related Health Problems - Tenth Revision (ICD-10) codes. Cardiovascular death was identified by codes: I00-I09, I11, I13, I20-I51. Other outcomes related to ASCVD, including nonfatal myocardial infarction and stroke, were not available in the NHANES III dataset. Death events were right-censored at 10 years. The full list of clinical variable names and their corresponding NHANES III codes are listed in **Supplemental Table 1**.

For ECG data, 133 features based on direct ECG measurements were used. Due to the high proportion of missing values, preprocessing steps for ECG data included removing rows (participants) and columns (ECG features) which had >50% missing data. Remaining missing values were imputed using the Multivariate Imputation by Chained Equations (*mice*) package^17^, based on demographic variables and other ECG features. A full list of selected ECG features is highlighted in **Supplemental Table 2**. Further preprocessing steps included converting the rhythm code to a binary variable (sinus vs. non-sinus rhythm) to avoid data sparsity and replacing QT interval with corrected QT interval based on Bazett’s formula. All ECG data were standardized to have a mean of 0 and standard deviation of 1.

Prior to model training, data were split into 80:20 train:test partitions by random sampling. Given the low frequency of outcome events, the training set was augmented by two procedures to improve class imbalance: 1) oversampling of positive events and 2) synthetic data generation using the Random Over-Sampling Examples (*ROSE*) package^18^, which creates a synthetic data sample of balanced class and parameter distribution by drawing new examples from a conditional kernel density estimate of the majority and minority classes.

### Model training

All machine learning models were trained via 10-fold cross validation on six different training data combinations based on the following schema: Three training data sets (base, oversample, synthetic) in two different parameter combinations (PCE + ECG variables or Demographic (age, sex, race) + ECG variables). Model performance was assessed in a single hold out test set, which was not used for any part of the model training process. As a comparison, the PCE was implemented as a Cox proportional hazards model based on published parameters^2,26^. For machine learning, various classification models were trained with 10-year cardiovascular mortality as a binary outcome. Logistic regression, random forest, gradient boosting machine, and support vector machine models were trained using the *caret*^19^ package with automated hyperparameter tuning, while for neural networks, the R implementation for *Keras*^20^ package was used for deep learning based on multilayer perceptron architecture with three hidden layers and regularization via dropout. Finally, ensemble models were trained using the *SuperLearner*^21^ package. Briefly, ensemble models compute multiple models (“base learners”) and aggregate their predictions, which improves overall accuracy but increases computational costs. For this study, ensemble models were built using logistic regression, random forest, gradient boosting machine, support vector machine, and neural network models as base learners, and optimal model weighting was determined by maximizing area under receiver operating characteristics curve (AUROC) based on the Nelder-Mead method and 5-fold cross validation.

### Model assessment and comparison

Discriminative performance of classification models and their ensembles were compared using AUROC, area under the precision-recall curve (AUPRC), sensitivity, and specificity metrics. To allow for comparison with the PCE, classification performance metrics were computed for the PCE by assessing the probability of cardiovascular death at or before 10 years, with threshold value set to maximize AUPRC. Machine learning models with superior prediction performance characteristics were further assessed using standard calibration plots.

To assess the prognostic value of individual ECG features, the variable importance rank metric from the *caret*^*19*^ package was compared, except for neural network and ensemble models where such variable importance rank metrics were not available. For aggregate assessment of classification models, the cumulative count of top ten important predictors for each model was counted and plotted on a standard 12-lead ECG for visual assessment and clinical interpretation.

## RESULTS

### Study population

Baseline characteristics of the study population are summarized in **Table 1**. The study cohort (N=7,067) had baseline age range of 40 to 90 years, with mean 59.2 and standard deviation of 13.4 years. Notably, while the initial inclusion criteria included all adults age 18 years and above, ECG data were only available for age 40 and above. For sex, there was a slight female majority (52.5%), while for race there was a significant majority in white (73.9%), followed by black (23.3%), and under-representation of other races. There was a wide range in body mass index (mean: 27.6, range: 13.3-64.5) and a significant comorbidity burden, including hypertension (33.8%), hyperlipidemia (23.3%), diabetes mellitus (46.7%), and tobacco use (24.5%). Follow-up period was 18.1±7.6 years, with 10-year all-cause mortality rate of 19.8% and cardiovascular mortality rate of 4.8%.

**Table 1.** Characteristics of the study population

| Category | Variable (units) | Mean ± SD [Range] or N (%) |
| --- | --- | --- |
| Demographics | Age (years) | 59.2 ± 13.4 [40.0-90.0] |
|  | Sex Male | 3,355 (47.5%) |
|  | Female | 3,712 (52.5%) |
|  | Race White | 5,223 (73.9%) |
|  | Black | 1,645 (23.3%) |
|  | Other | 199 (2.8%) |
| Vital signs | Heart rate (beats/min) | 74 ± 10 [43-164] |
| | Systolic BP (mmHg) | 132 ± 20 [78-248] |
| | Diastolic BP (mmHg) | 77 ± 10 [16-136] |
| Body measurements | Weight (kg) | 76.2 ± 17.1 [33.4-182.3] |
| | Height (cm) | 165.9 ± 9.9 [126.9-200.0] |
| | Body mass index (kg/m<sup>2</sup>) | 27.6 ± 5.5 [13.3-64.5] |
| Medical history | Hypertension | 2,385 (33.8%) |
|  | Hyperlipidemia | 1,645 (23.3%) |
|  | Diabetes mellitus | 3,297 (46.7%) |
|  | Tobacco use (current) | 1,732 (24.5%) |
| Medications | Medication for blood pressure | 2,238 (31.7%) |
|  | Medication for cholesterol | 203 (2.9%) |
| Laboratory | Total cholesterol (mg/dL) | 217 ± 43 [59-501] |
| | HDL cholesterol (mg/dL) | 51 ± 16 [12-196] |
| | LDL cholesterol (mg/dL)<sup>a</sup> | 136 ± 38 [20-361] |
| | Triglycerides (mg/dL) | 157 ± 116 [27-3616] |
| | HgbA1c (%) | 5.8 ± 1.2 [2.7-16.1] |
| | C-reactive protein (mg/dL) | 0.5 ± 0.8 [0.2-18.3] |
| Follow-up | Follow-up (years) | 18.1 ± 7.6 [0.0-27.2] |
| Outcomes | 10-year all-cause mortality | 1,399 (19.8%) |
|  | 10-year cardiac mortality | 338 (4.8%) |
|  | 10-year cerebrovascular mortality | 111 (1.6%) |

SD: Standard deviation; <sup>a</sup>significant missing proportion (55.7%)

### Prediction performance of machine learning models

Best performing machine learning models and their classification metrics are shown in **Table 2**. Compared to the PCE (AUROC: 0.668 [95% confidence interval: 0.625-0.712], AUPRC: 0.125 [0.081-0.170], sensitivity: 0.492, specificity: 0.859), several machine learning models trained on the PCE + ECG variable combination showed improved performance: logistic regression (AUROC: 0.762 [0.723-0.800], AUPRC: 0.143 [0.100-0.185], sensitivity: 0.759, specificity: 0.756), neural network (AUROC: 0.729 [0.646-0.813], AUPRC: 0.153 [0.089-0.216], sensitivity: 0.658, specificity: 0.822), and ensemble model (AUROC: 0.683 [0.648-0.718], AUPRC: 0.146 [0.092-0.199], sensitivity: 0.494, specificity: 0.884). Limiting training data to only demographic and ECG variables did not adversely affect predictive performance of machine learning models: logistic regression (AUROC: 0.754 [95% CI: 0.702-0.806], AUPRC: 0.141 [0.093-0.189], sensitivity: 0.747, specificity: 0.759), neural network (AUROC: 0.764 [0.712-0.815], AUPRC: 0.149 [0.110-0.188], sensitivity: 0.722, specificity: 0.787), ensemble model (AUROC: 0.695 [0.668-0.722], AUPRC: 0.166 [0.116-0.215], sensitivity: 0.468, specificity: 0.912). Comparison plots of AUROC and AUPRC are shown in **Figure 2**. When predicted results were plotted as survival curves, there was clear separation between those predicted to be with and without cardiovascular death occurring at or before 10 years, with the best curve separation achieved by ensemble models (**Figure 3**).

**Table 2.** Model performance comparison

| **Model based on PCE + ECG data** | **AUROC [95% CI]** | **AUPRC [95% CI]** | **Sensitivity** | **Specificity** |
| --- | --- | --- | --- | --- |
| Pooled Cohort Equations<sup>a</sup> | 0.668 [0.625-0.712] | 0.125 [0.081-0.170] | 0.492 | 0.859 |
| Logistic regression<sup>b</sup> | 0.762 [0.723-0.800] | 0.143 [0.100-0.185] | 0.759 | 0.756 |
| Neural network<sup>c</sup> | 0.729 [0.646-0.813] | 0.153 [0.089-0.216] | 0.658 | 0.822 |
| Ensemble model<sup>c</sup> | 0.683 [0.648-0.718] | 0.146 [0.092-0.199] | 0.494 | 0.884 |
| **Model based on demographic + ECG data** | **AUROC [95% CI]** | **AUPRC [95% CI]** | **Sensitivity** | **Specificity** |
| Pooled Cohort Equations<sup>a</sup> | 0.668 [0.625-0.712] | 0.125 [0.081-0.170] | 0.492 | 0.859 |
| Logistic regression<sup>b</sup> | 0.754 [0.702-0.806] | 0.141 [0.093-0.189] | 0.747 | 0.759 |
| Neural network<sup>c</sup> | 0.764 [0.712-0.815] | 0.149 [0.110-0.188] | 0.722 | 0.787 |
| Ensemble model<sup>c</sup> | 0.695 [0.668-0.722] | 0.166 [0.116-0.215] | 0.468 | 0.912 |

AUPRC: Area under precision-recall curve, AUROC: Area under receiver operating characteristic curve, CI: Confidence interval (based on 5-fold cross validation), ECG:
Electrocardiogram, PCE: Pooled Cohort Equations
<sup>a</sup>Classification performance assessed at 10 years, with threshold value set to maximize AUPRC
<sup>b</sup>model trained on oversampled training set
<sup>c</sup>model trained on synthetic training set

**Figure 2.**
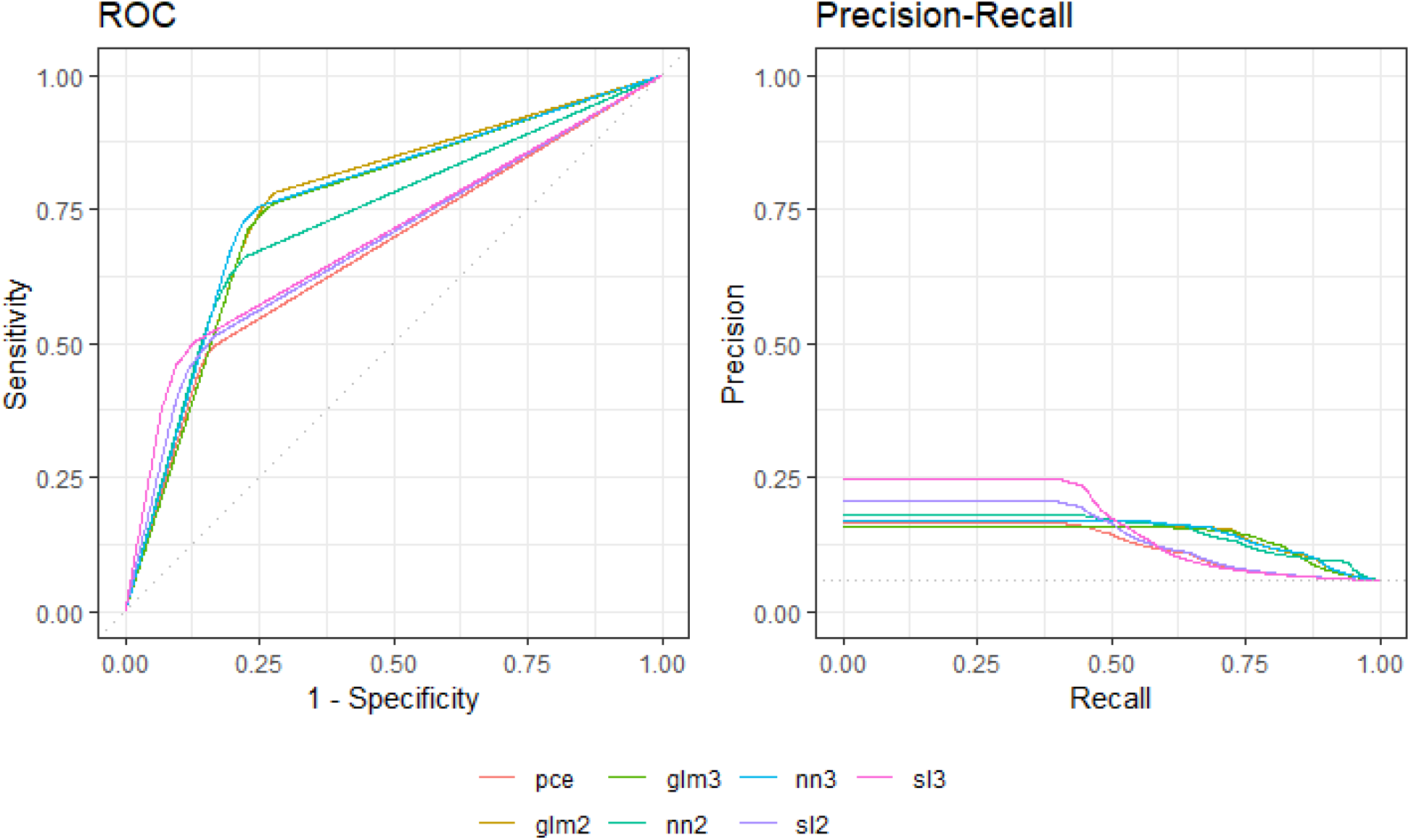
Receiver operator characteristic (ROC) and Prediction-recall curves of machine learning models glm2: Logistic regression (PCE+ECG data), glm3: Logistic regression (Demographic+ECG data), nn2: Neural network (PCE+ECG data), nn3: Neural network (Demographic+ECG data), pce: Pooled Cohort Equations, sl2: Ensemble model (PCE+ECG data), sl3: Ensemble model (Demographic+ECG data)

**Figure 3.**
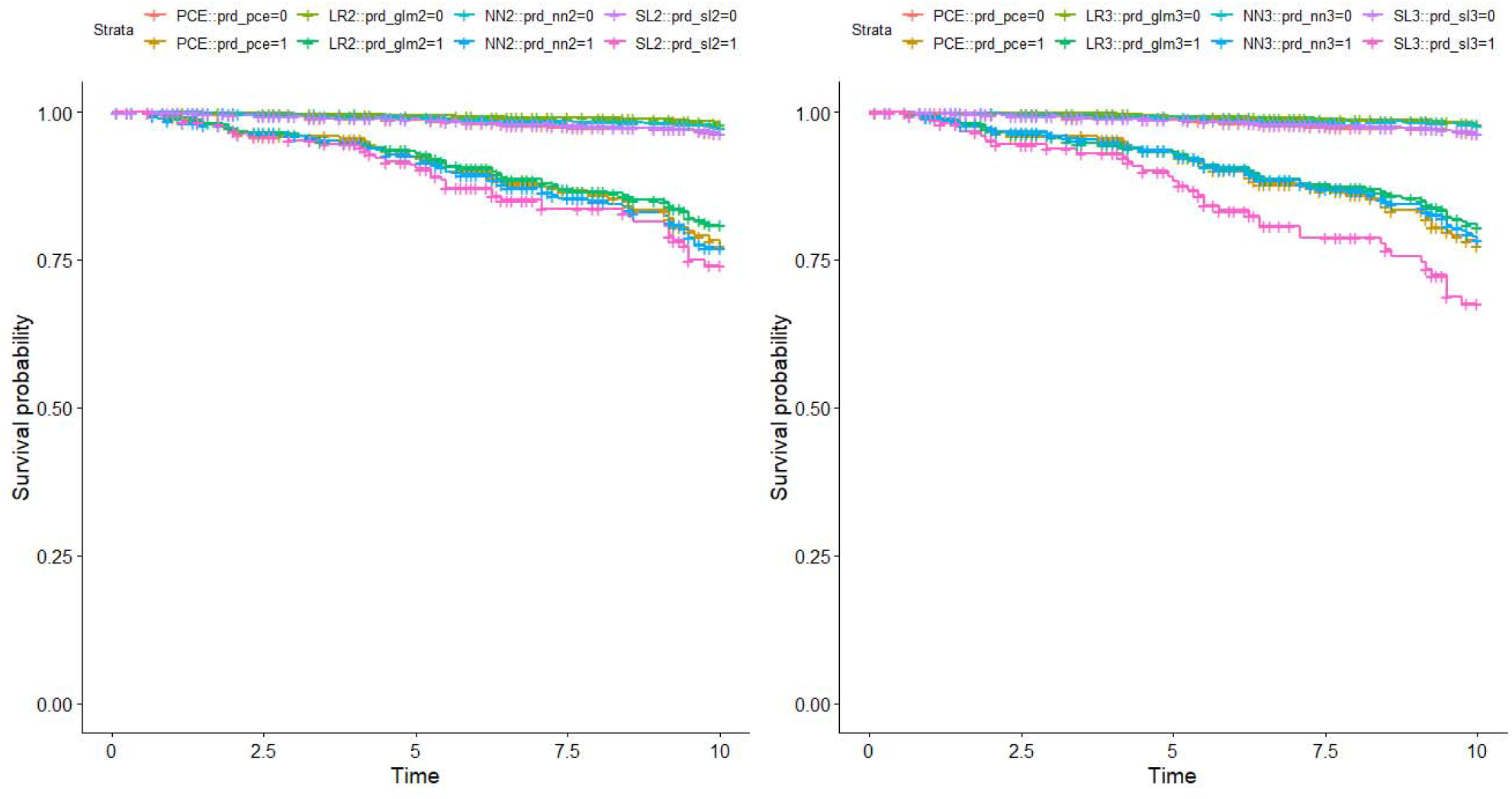
Survival curves based on model prediction (Left-side): Models based on PCE+ECG data. pce: Pooled Cohort Equations, glm2: Logistic regression, nn2: Neural network, sl2: Ensemble model (Right-side): Models based on Demographic+ECG data. pce: Pooled Cohort Equations, glm3: Logistic regression, nn3: Neural network, sl3: Ensemble model

Training data augmentation generally had beneficial effects on model classification performance, though the degree varied by model family (**Supplemental Table 3**). For logistic regression, both oversampled and synthetic training data markedly improved classification performance. For neural networks, there was sequential improvement in performance from base to oversampled to synthetic training data. For ensemble models, base training data worked well, with only comparable performance seen with synthetic data. Training data augmentation also generally improved model calibration (**Supplemental Figure 1**). In general, models tended to overestimate risk when predicting higher event probability. Calibration curves were haphazard for base logistic regression and ensemble models, compared to smoother calibration curves for models based on oversampled and synthetic data, suggesting improved calibration when using augmented training data. However, training data augmentation did not improve classification performance or calibration of random forest, gradient boosting machine, and support vector machine models, which all had poor predictive value overall.

### Variable importance comparison

Among traditional cardiovascular risk factors, age was the most important predictor of 10-year cardiovascular mortality, occurring as the top variable in all models, followed by systolic blood pressure and treatment for hypertension. Among ECG features, the most important were R amplitude in lead II, J amplitude in lead V5, J amplitude in lead V6, R amplitude in lead aVL, S amplitude in lead III, S amplitude in lead aVF, and S duration in lead V6. Interestingly, the most important ECG features appeared to cluster in inferior (II, III, aVF) and lateral (aVL, V5, V6) leads, as easily seen when plotted on a standard 12-lead ECG (**Figure 4**). Ranking of important predictor variables for individual models and the aggregate counts for the top ten predictors for each model are shown in **Supplemental Table 4**.

**Figure 4.**
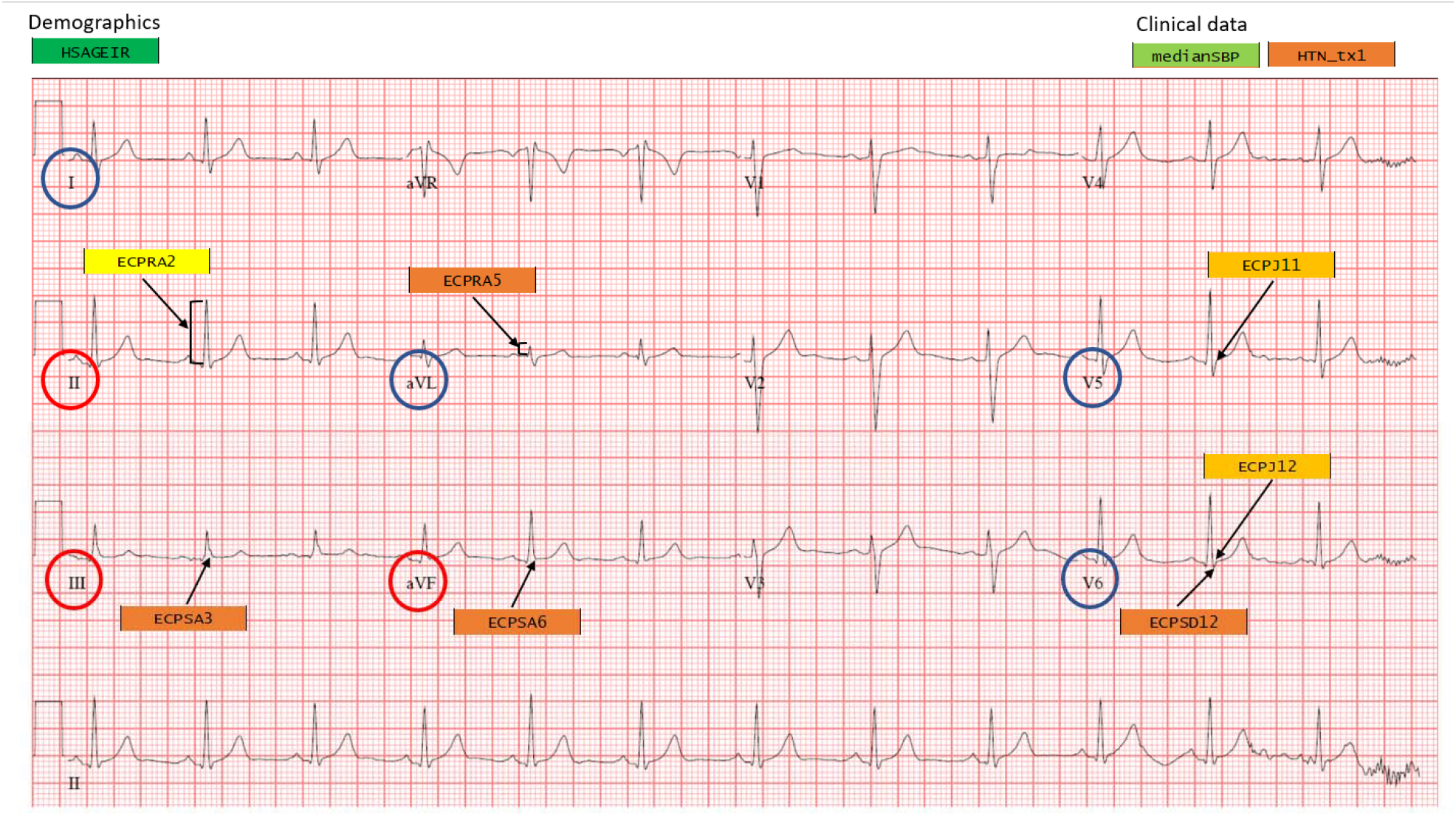
Variable importance plot on the 12-lead electrocardiogram Color schema is based on decreasing frequency of occurrence in machine learning models: Dark green (HSAGEIR: Age), Light green (medianSBP: Systolic blood pressure), Yellow (ECPRA2: R amplitude, lead II), Light orange (ECPJ11: J amplitude, lead V5; ECPJ12: J amplitude, lead V6), Orange (HTN_tx1: Treatment for hypertension; ECPRA5: R amplitude, lead aVL; ECPSA3: S amplitude, lead III; ECPSA6: S amplitude, lead aVF; ECPSD12: S duration, lead V6). Red and blue circles indicate inferior and lateral lead groupings of the 12-lead ECG, respectively.

## DISCUSSION

In this study, we showed that machine learning methods could be pragmatically applied to aggregate ECG data to predict 10-year cardiovascular mortality. Machine learning models, particularly the logistic regression, neural network, and ensemble models, performed favorably in terms of classification metrics compared to the current clinical standard, the PCE. Only demographic and ECG features were required to achieve comparable performance, without requiring traditional cardiovascular risk factors represented in the PCE. Interestingly, individual ECG features with the most prognostic information were seen to cluster in inferior and lateral segments of the ECG.

Prior studies have examined individual ECG components (e.g. P wave duration^4^, deep terminal negativity of P wave in V1^5,27^, QRS duration^6^, QT interval^28^, JT interval^29^, and isolated ST-segment and T-wave abnormalities^30^) or groups of ECG components^31^ for their moderate additive predictive value with respect to standardized cardiovascular risk calculators such as the Framingham Risk Score or the PCE. Beyond individual components, groups of ECG components have been evaluated in the framework of global electrical heterogeneity^32^, though this is not typically computed or utilized in clinical practice. Despite the potential, the United States Preventive Services Task Force in 2018 recommended against using the ECG to screen adults with low risk of cardiovascular events (grade D), and remained undecided for adults with presumed intermediate or high risk of cardiovascular events (grade I), citing the imbalance between the potential benefits of early disease detection versus harms related to unnecessary invasive testing and overtreatment^33^.

Subsequent studies employing modern machine learning techniques, however, have shown that aggregate ECG data contain significant predictive information for detection of various cardiac abnormalities such as systolic dysfunction^10,34^, diastolic dysfunction^9^, atrial fibrillation^12^, pulmonary arterial hypertension^8^, hypertrophic cardiomyopathy^8^, cardiac amyloid^8^, mitral valve prolapse^8^, as well as prognostic information for short term mortality^35^. Expanding on this trend, this study has demonstrated that machine learning models based on aggregate ECG data can also predict long-term outcomes such as 10-year cardiovascular mortality. In fact, demographic and ECG features appear to contain as much prognostic information as rest of traditional cardiovascular risk factors represented in the PCE, with potentially important implications for primary prevention such as obviating the need to obtain blood draws for long-term cardiovascular risk prediction.

The important individual ECG features that contributed to model performance were not the traditional markers of ischemia or infarction (e.g. abnormalities in Q wave, ST segment, or R wave progression) that would be expected on a clinical basis. These are supportive of the findings of a recent study by Raghunath *et al*.^35^, where subclinical ECG markers were most predictive of short-term mortality. While it is conceivable that a significant portion of the study population had clinically silent coronary artery disease that manifested in the inferior and lateral leads of the ECG, these findings could also represent an entirely different mechanism that contributes to long-term cardiovascular mortality risk, or simply highly correlated data. Further studies are needed to validate these findings.

### Limitations

There are several important limitations to this study. First, this was a retrospective study based on a single data source, with the usual limitations associated with this study design. Not all data components, specifically ECG data, were available for all participants of NHANES III. Therefore only a subset of the survey participants formed the study cohort, who may not be representative of the target population of undifferentiated ambulatory adults. There was a significant proportion of missing values in ECG data, requiring substantial preprocessing steps and data imputation. It is possible that the ECG data were not missing at random, and possibly impacted by variable lead placement techniques, which may have led to biased results. Machine learning models were trained as classification and not survival models, which can lead biased parameter estimates. However, parameter estimation was not a focus of this study and survival curves based on model prediction showed clear curve separation. Finally, some machine learning models were not at all effective for event prediction in test data, revealing the potential for overfitting in any machine learning algorithm. Despite these limitations, the comparative analysis framework adopted in this study with less focus on individual model parameters but greater emphasis on aggregate findings demonstrated the feasibility and utility of applying machine learning to aggregate ECG data for prediction of long-term cardiovascular mortality.

## CONCLUSIONS

Machine learning can be applied to demographic and ECG data to predict 10-year cardiovascular mortality in ambulatory adults, with potentially important implications for primary prevention of cardiovascular disease. Further studies are needed to validate these findings.

## Data Availability

NHANES III is a publicly available, de-identified data set.

https://www.n.cdc.gov/nchs/nhanes/nhanes3/default.aspx

## ABBREVIATIONS

ASCVD: Atherosclerotic cardiovascular disease
AUPRC: Area under precision-recall curve
AUROC: Area under receiver operating characteristics curve
ECG: Electrocardiogram
NHANES: National Health and Nutrition Examination Survey
PCE: Pooled Cohort Equations

## ACKNOWLEDGEMENTS

The primary author would like to express gratitude to Dr. Jarrod Dalton, PhD at Cleveland Clinic, who provided the R code adaptation for the 2013 ACC/AHA Pooled Cohort Equations via personal communication.

