## Supplemental material for "Machine Learning to Predict 10-year Cardiovascular Mortality from the Electrocardiogram: Analysis of the Third National Health and Nutrition Examination Survey (NHANES III)"

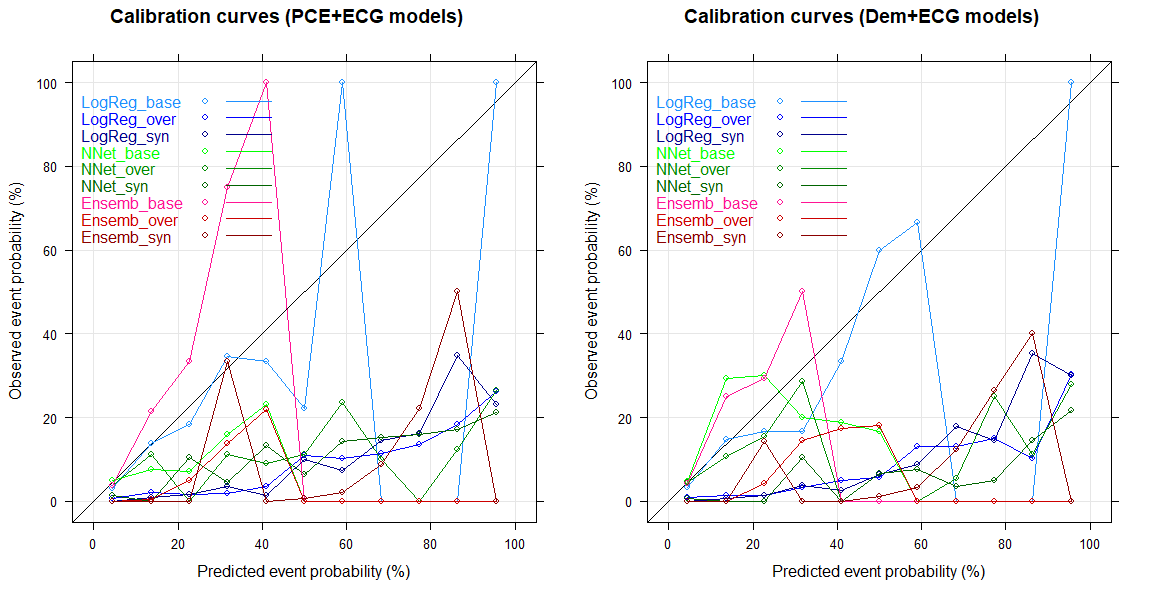

**Supplemental Figure 1**. Calibration curves of machine learning models.

LogReg: Logistic regression

Nnet: Neural network

Ensemb: Ensemble model

base: model trained on base training set

over: model trained on oversampled training set

syn: model trained on synthetic training set

**Supplemental Table 1**. Demographic and clinical variables from NHANES III.

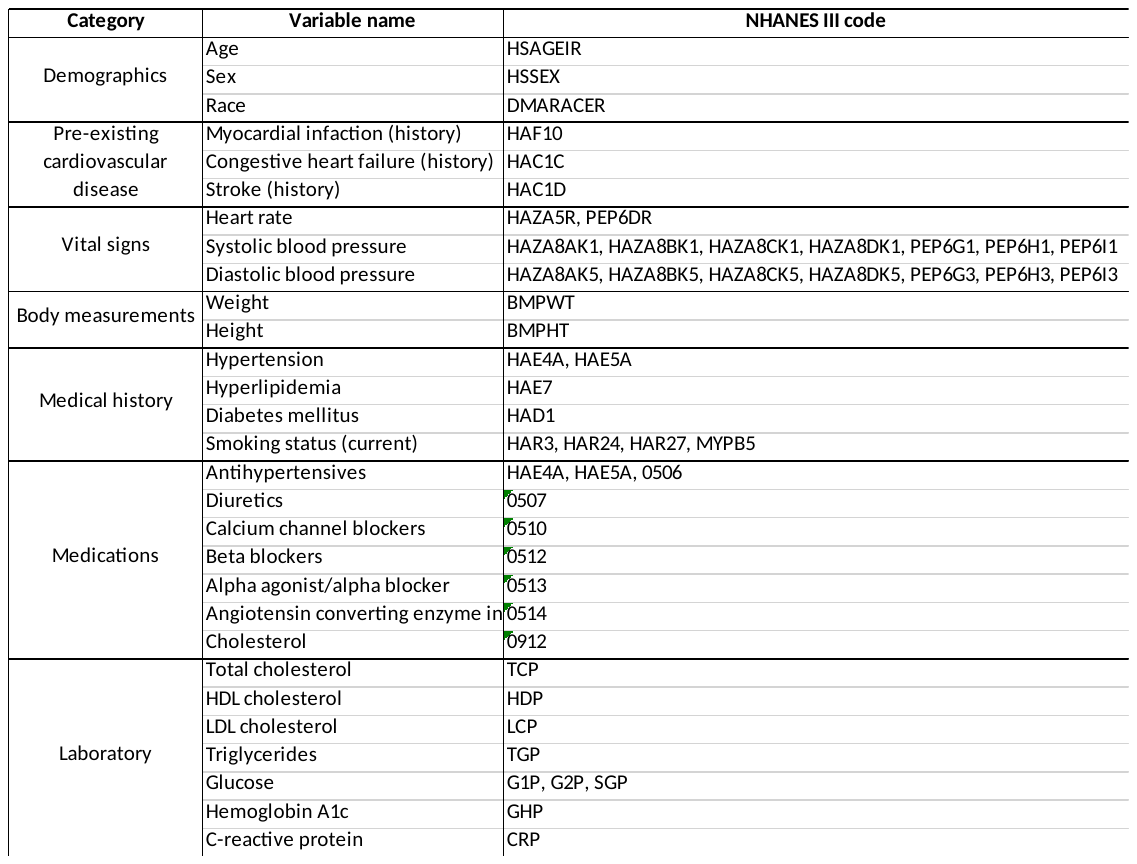

**Supplemental Table 2**. Selected electrocardiogram measures from NHANES III (highlighted variables used for model training following pre-processing)

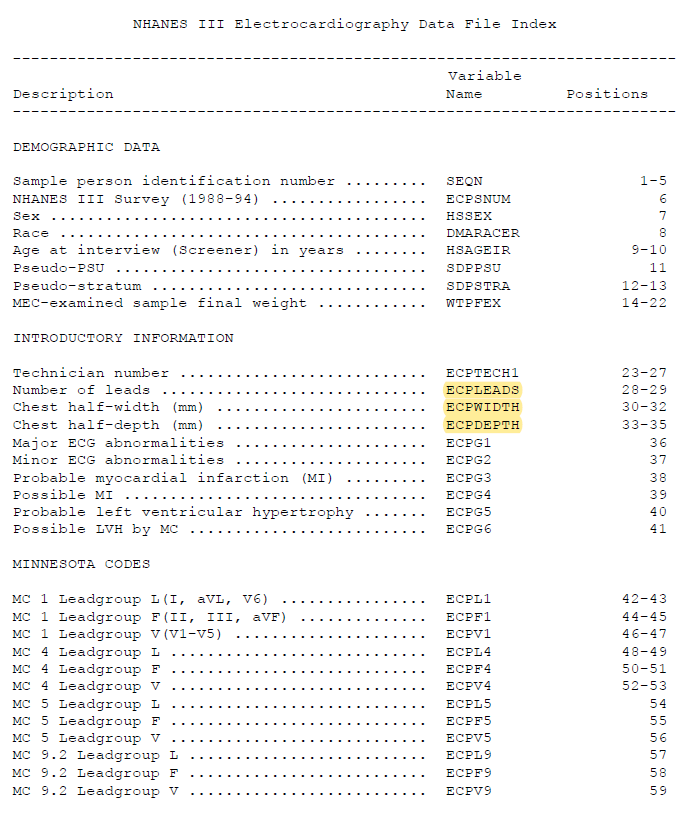

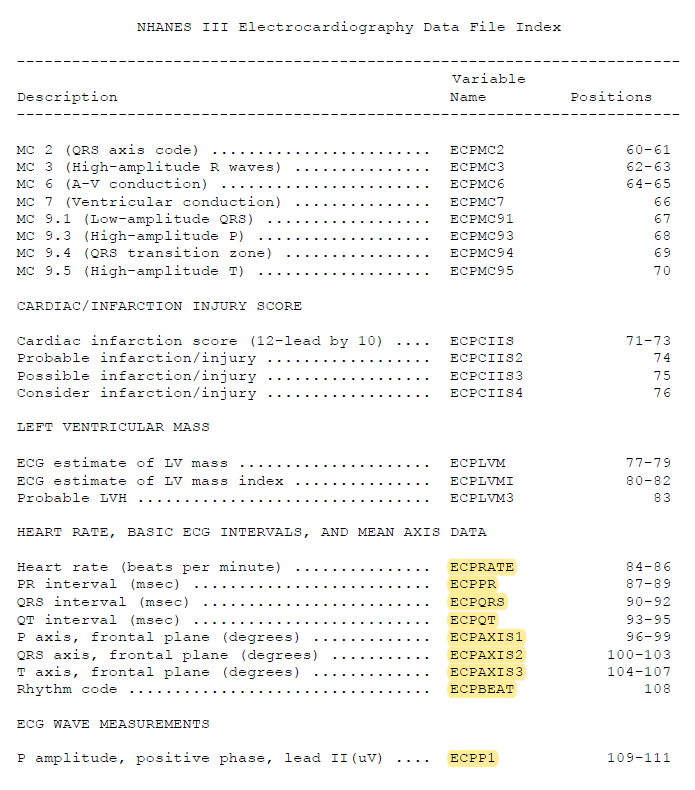

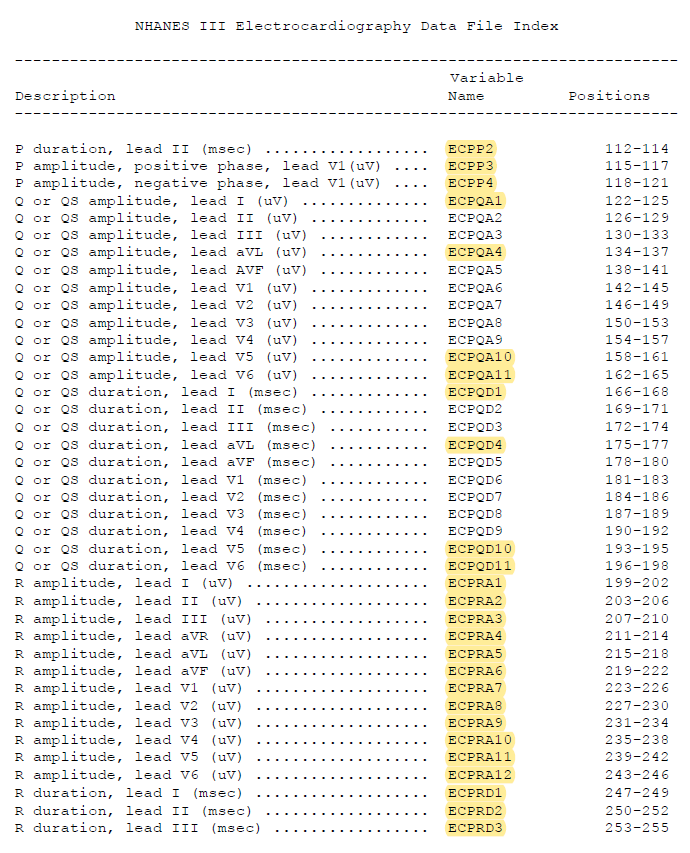

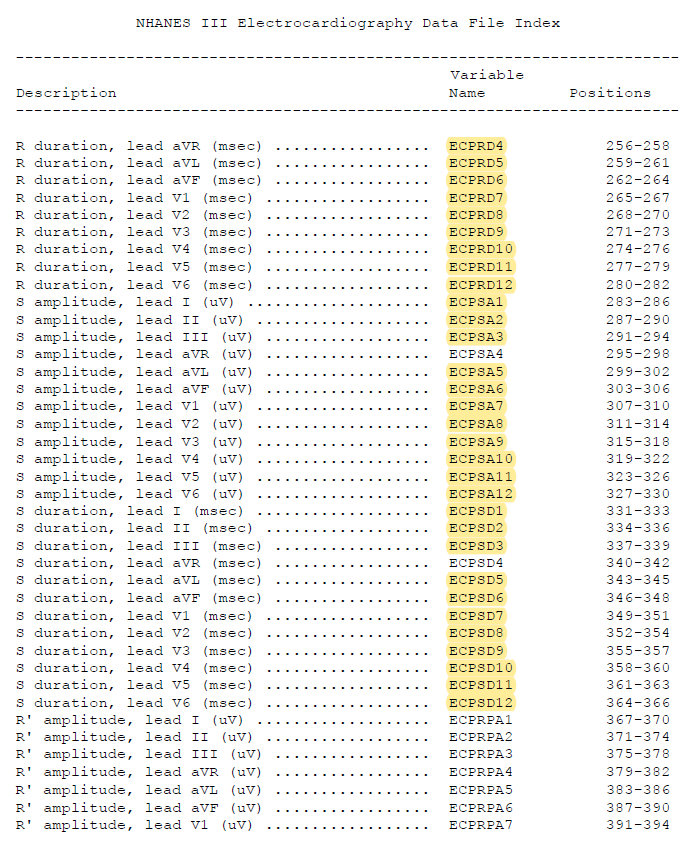

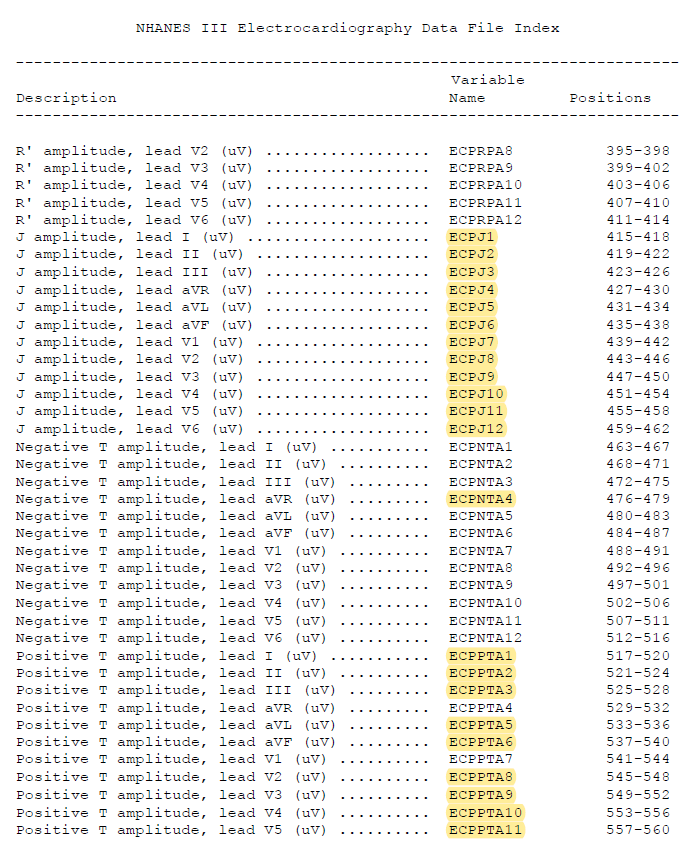

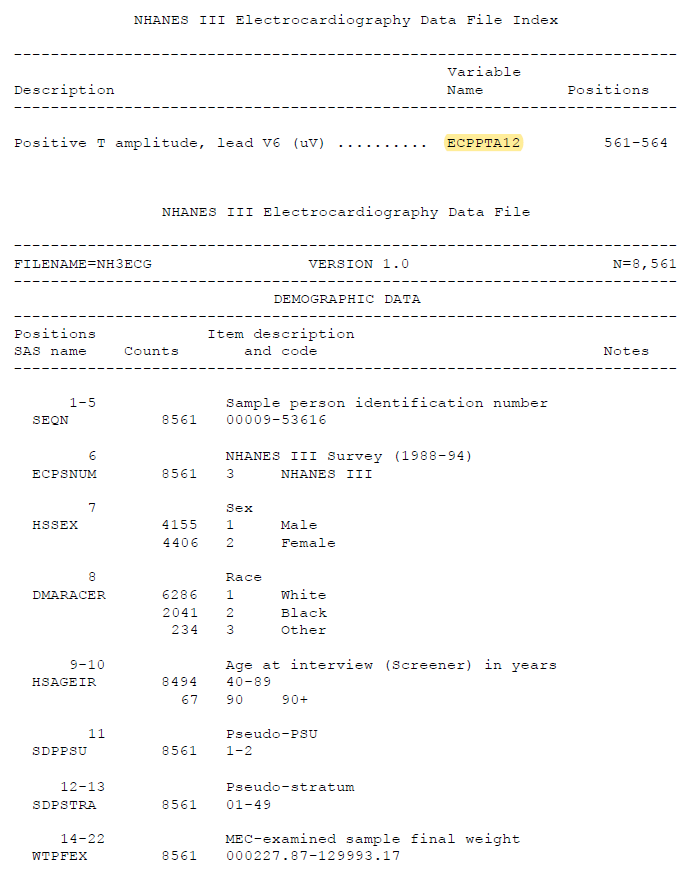

**Supplemental Table 3**. Model performance comparison (full).

PCE: Pooled Cohort Equations, ECG: Electrocardiogram, AUROC: Area under receiver operating characteristic curve, AUPRC: Area under precision-recall curve, Confidence Interval (bootstrap)

^a^Classification performance assessed at 10 years, with threshold value set to maximize AUPRC

^b^model trained on base training set

^c^model trained on oversampled training set

^d^model trained on synthetic training set

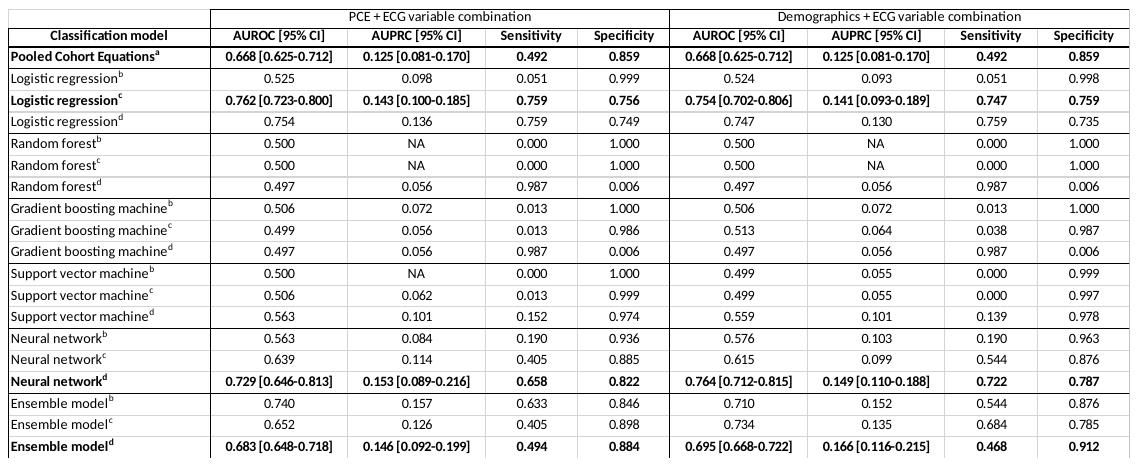

**Supplemental Table 4**. Variable importance comparison between prediction models

LR: Logistic regression, GBM: Gradient boosting machine, SVM: Support vector machine

^a^model trained on base training set

^b^model trained on oversampled training set

^c^model trained on synthetic training set

Color schema based on frequency of importance: Dark green(7), Light green(6), Yellow(5), Light orange(4), Orange(3)

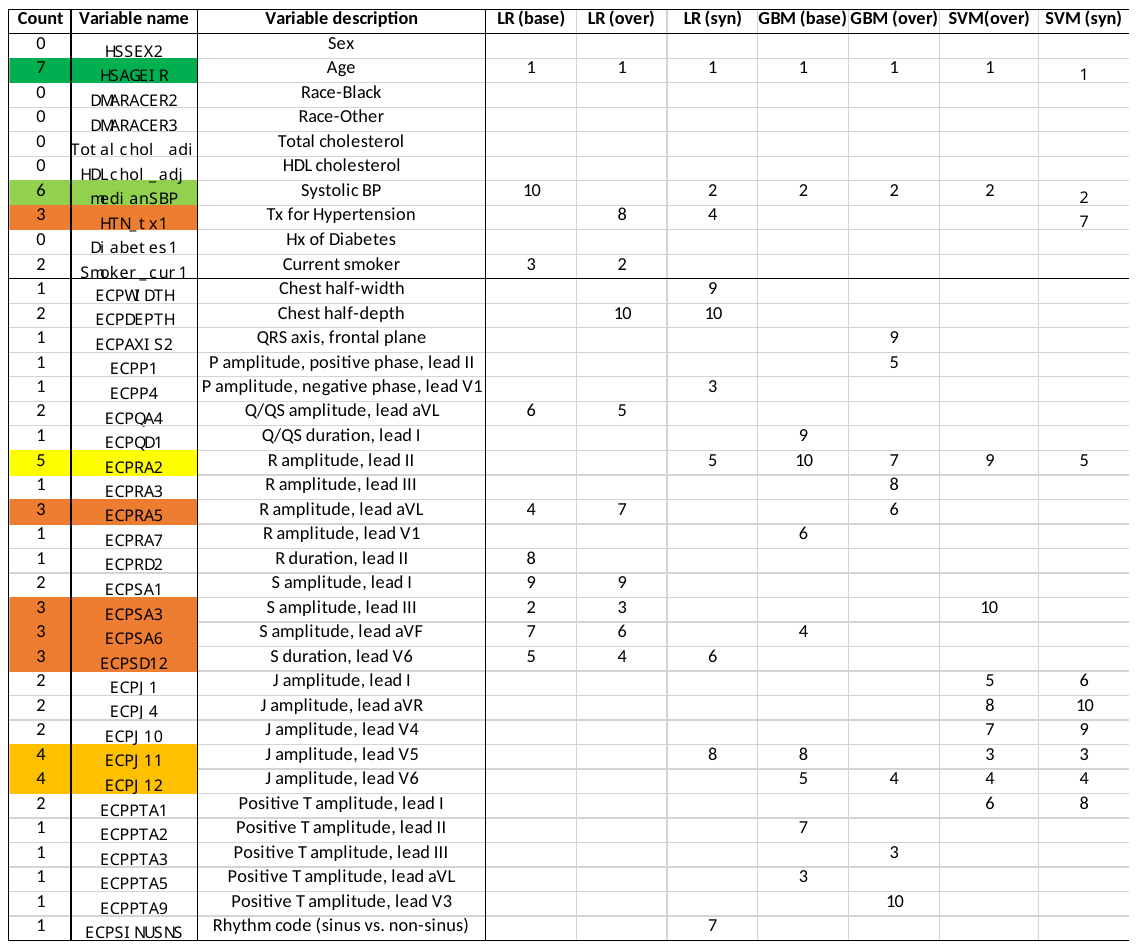
